# Effect of five years of biannual azithromycin mass drug administration on enteric fever seroincidence in Niger: A randomized control trial

**DOI:** 10.64898/2026.08.03.26359544

**Authors:** Kristen Aiemjoy, Jessica C Seidman, Ahmed M Arzika, Ramatou Maliki, Amza Abdou, Douglas Ezra Morrison, Kristina W Lai, Denise O. Garrett, Claire Munroe, Abel Gonzalez, Leah Sukri, Elodie Lebas, Catherine Cook, Benjamin F Arnold, Thomas M Lietman, Kathleen M Neuzil, Jason R Andrews, Jeremy D Keenan, Richelle C Charles

## Abstract

**Background:** Enteric fever, caused by *Salmonella enterica* serovars Typhi and Paratyphi, remains a major cause of morbidity in low- and middle-income countries, yet burden estimation is challenging in regions without blood culture surveillance. Azithromycin is effective for treating enteric fever, and mass drug administration (MDA) to young children reduces all-cause mortality in sub-Saharan Africa, but its impact on community-level enteric fever burden is unknown. We assessed enteric fever seroincidence in rural Niger and evaluated the effect of biannual azithromycin MDA affected.

**Principal Findings:** This was a secondary analysis of the MORDOR trial (NCT02048007), a cluster-randomized, placebo-controlled study in 30 communities in Dosso Region, Niger. Children aged 1–59 months received biannual azithromycin or placebo. Dried blood spots collected at baseline (2015) and Year 5 (2020) were tested for IgA and IgG antibodies against Hemolysin E using kinetic ELISA. Seroincidence was estimated using maximum likelihood methods based on antibody kinetics from blood culture-confirmed enteric fever patients. Samples from 1,417 children were analyzed (423 baseline; 994 Year 5). Overall seroincidence approximately doubled from baseline to Year 5, increasing from 68.5 (95% CI 58.0–80.7) to 145.2 (95% CI 132.3–159.3) per 100 person-years in placebo communities and from 74.7 (95% CI 56.5–98.7) to 135.2 (95% CI 112.4–162.6) in azithromycin communities, with no significant difference between arms (difference-in-differences −16.2; 95% CI -53.1-20.8 ). Among children <2 years, the increase in seroincidence was smaller in the azithromycin arm (37.5; 95% CI: 1.3-73.6) compared to placebo (81.1; 95% CI 54.0-108.2), with a difference-in-differences of −43.7 per 100 person-years (95% CI −88.8 - 1.5 to 0.02).

**Significance:** Enteric fever seroincidence among young children in rural Niger is high and increased substantially over the five-year study period. Biannual azithromycin MDA did not significantly reduce overall seroincidence, though a potential attenuation was observed among children under 2 years of age. These findings underscore the importance of Niger’s recent typhoid conjugate vaccine introduction and highlight the value of serosurveillance for monitoring enteric fever infection intensity in settings without blood culture surveillance.

## BACKGROUND

Enteric fever, caused by *Salmonella enterica* serovars Typhi and Paratyphi, remains a significant public health concern in low- and middle-income countries, with an estimated 14 million cases and over 100,000 deaths globally each year.^1^ The burden is concentrated in South Asia and sub-Saharan Africa, where limited access to safe water, sanitation, and hygiene infrastructure perpetuates transmission.^1^ Accurate surveillance for enteric fever is hampered by the lack of affordable and accurate diagnostics, particularly in resource-limited settings.^2,3^ Blood culture, the reference standard for diagnosis, requires expensive laboratory infrastructure, trained personnel, and continuous supply chains for culture media and consumables, making systematic surveillance prohibitively costly in many low-income settings. In sub-Saharan Africa, blood culture surveillance data for enteric fever remain scarce, hindering accurate burden estimation and impeding evidence-based vaccine introduction and other preventive interventions.^4,5^

Niger faces a particularly high burden of enteric infections among young children. The country has one of the highest rates of diarrheal disease mortality globally, with an estimated 485 deaths per 100,000 children under 5 years in 2015 when the baseline study visit was conducted.^6^ Although blood culture–confirmed data for enteric fever are limited, surgical case series provide indirect evidence of extensive infection burden. Typhoid intestinal perforation (TIP)—a late complication of untreated *S.* Typhi infection—results from an immune-mediated hypersensitivity reaction (Shwartzman/Koch-type) at the Peyer’s patches, which may be more severe following prior sensitization. TIP has been reported at exceptionally high levels in Niger. A recent scoping review across 12 Francophone African countries found Niger to have the highest clinically-diagnosed TIP caseload, including one study from Zinder and Maradi that reported 2,931 cases between 2014 and 2019 ^7,8^. The median patient age was 11 years (IQR 7–17), with children comprising more than 70% of cases.^8^ This age pattern is notable, as TIP typically affects older adolescents and young adults.^9^ The predominance of TIP in younger children in Niger may reflect high rates of exposure early in life—consistent with intense transmission in this population. Because perforation occurs in only 1–3% of untreated typhoid cases, these reports imply a large underlying incidence of infection.^10^ However, systematic blood culture surveillance has not been established in Niger, leaving the true burden of disease unquantified and highlighting the need for scalable methods to estimate infection burden.

Azithromycin is a broad-spectrum antibiotic that effectively treats uncomplicated enteric fever and is recommended by WHO as a first-line therapy, particularly in settings with fluoroquinolone resistance^11^. Beyond its role in treating clinical disease, mass drug administration (MDA) of azithromycin to young children has been shown to reduce all-cause mortality by 13.5% in sub-Saharan Africa, with the largest reduction observed in Niger of 18.1%^12,13^. Although the exact mechanism of this mortality reduction remains unclear, it is thought to involve decreased deaths due to malaria, pneumonia, and diarrhea—raising the possibility that azithromycin MDA may also reduce infection with enteric pathogens, including *S.* Typhi^14,15^. However, the widespread use of azithromycin raises concerns about antimicrobial resistance. Azithromycin resistance in *S.* Typhi and *S.* Paratyphi A has emerged in South Asia since 2013, mediated by a single point mutation (R717Q/L) in the *acrB* gene encoding an efflux pump^16,17^. While azithromycin resistance remains relatively uncommon (0.6-2% in most South Asian settings), its spontaneous emergence in multiple independent lineages raises concerns about the potential for spread, particularly with increasing azithromycin use^18,19^. The impact of mass azithromycin distribution on enteric fever incidence and the potential for selection of resistant strains remains unknown.

Novel serological methods now enable estimation of enteric fever incidence from cross-sectional serosurveys in settings lacking blood culture surveillance^20,21^. These approaches measure quantitative IgA and IgG antibodies to Hemolysin E (HlyE), a pore-forming toxin secreted by *S.* Typhi and *S.* Paratyphi A^22^. By coupling these specific serologic markers with modeled antibody decay trajectories from blood culture-confirmed enteric fever patients, these methods estimate the seroincidence rate (the rate at which new infectious occur in a population) from cross-sectional antibody distributions. Validation studies in South Asia have demonstrated that serosurvey-based incidence estimates correlate with the rank order of burden observed in blood culture surveillance^20^. This methodology has successfully identified high forces of infection in settings without blood culture surveillance, including South Sudan and Kenya, where serosurvey data revealed substantial number of incident enteric fever infections that had not been previously documented^23,24^. This study aimed to apply these methods to assess enteric fever seroincidence in rural Niger and to evaluate whether biannual azithromycin mass drug administration impacted enteric fever seroincidence rates.

## METHODS

### Study Design and Setting

This was a secondary analysis of samples collected during a cluster-randomized, placebo-controlled study conducted in 30 rural communities in the Dosso region of Niger (NCT02048007).^25,26^ The trial was run concurrently with, and offered the same interventions as, the larger Macrolides Oraux pour Réduire les Décès avec un Oeil sur la Résistance (MORDOR) trial, which was designed to evaluate the effect of biannual mass azithromycin distributions on child mortality in sub-Saharan Africa (ClinicalTrials.gov, NCT02047981, https://clinicaltrials.gov/study/NCT03338244). Communities were randomly allocated in a 1:1 ratio to receive either azithromycin (approximately 20 mg/kg oral suspension) or placebo administered twice yearly. Randomization was performed at the community level using a computer-generated randomization scheme. In the present analysis, we used serological markers from stored dried capillary blood spot (DBS) samples to estimate enteric fever seroincidence. Details of the trial design, randomization procedures, and primary outcomes have been published previously.^25^

### Study Population and Eligibility

All children aged 1–59 months weighing >3.8 kg residing in the 30 study communities were eligible for treatment. For the annual serological surveys, a random sample of 40 children from each community was selected to provide capillary blood samples, with separate random samples selected at each annual monitoring visit based on the most recent biannual door-to-door study census (cross-sectional random samples). The present analysis used baseline (pre-treatment) samples collected from 11 March 2015 through 15 June 2015 and year 5 (month 60) samples collected from 4 February 2020 through 22 March 2020, following 10 rounds of biannual treatment. 15 DBS samples per community were randomly selected for testing from the baseline visit and all available samples were tested for the year 5 visit.

### Sample Collection and Storage

DBS were collected onto calibrated filter paper (TropBio Pty Ltd., Townsville, Queensland, Australia) and stored in individual sealable plastic bags with desiccant at −20°C until testing, except during shipment which occurred at ambient temperatures.

### Laboratory Methods

DBS were tested for enteric fever Hemolysin E (HlyE) antibodies as previously described.^20^ Briefly, DBS were eluted into phosphate-buffered saline with 0.05% Tween 20 prior to analysis. Plates were coated with purified HlyE (1 µg/mL). Eluates were loaded in duplicate and bound antibody was detected with goat anti-human IgG and IgA conjugated with horseradish peroxidase (Jackson ImmunoResearch), and peroxidase activity was measured at 450 nm using the chromogenic substrate, o-phenylenediamine. To compare across ELISA plates, the blank-adjusted sample readings were averaged, divided by the readings of a standard included on each plate (human chimeric monoclonal antibody for HlyE), multiplied by 100 and reported as K-ELISA units.

### Statistical Analysis

Seroincidence rates were estimated using the serocalculator package in R (1.4.0.9004), which implements maximum likelihood estimation of infection rates from cross-sectional antibody response data based on age-specific antibody kinetics.^20,27,28^ This approach models the probability of observing a given antibody level in an individual of age as a function of: (1) the force of infection (λ), (2) peak antibody levels post-infection, (3) the rate of antibody decay over time, and (4) measurement noise.

The analysis incorporated antibody decay trajectories from children under 5 years derived from the SeroEpidemiology and Environmental Surveillance (SEES) study, which followed blood culture-confirmed enteric fever patients in Nepal, Pakistan, and Bangladesh with longitudinal antibody measurements.^20^ These decay trajectories were modeled using two-phase power-function decay models fitted to longitudinal antibody responses using Bayesian hierarchical methods, accounting for individual variability in peak antibody levels and decay rates.

Seroincidence estimates were calculated using IgA antibody responses to HlyE in the primary analysis given recent evidence suggesting IgA responses and decay rates are more comparable across setting switch difference forces of infection.^29^ Seroincidence estimates using IgG are reported in the supplemental material.

Seroincidence was estimated separately for the baseline survey (2015) and Year 5 survey (2020), stratified by treatment group (azithromycin vs. placebo). Maximum likelihood estimates and 95% confidence intervals were calculated using profile likelihood methods with cluster-robust standard errors to account for the clustered sampling design. Estimates were further stratified by age groups of interest for typhoid vaccine policy (<2 years; 2–4 years).

The effect of azithromycin MDA on enteric fever seroincidence was assessed using a difference-in-differences approach, comparing the change in seroincidence from baseline to Year 5 between treatment arms. Standard errors for difference-in-differences estimates were calculated by propagation of uncertainty from the cluster-robust standard errors of the four component seroincidence estimates, treating estimates from baseline and Year 5 cross-sectional surveys as independent. Confidence intervals were calculated as the estimate ± 1.96 times the propagated standard error. Missing data we treated as missing at random (MAR).

Statistical analyses were conducted using R statistical software (version 4.3.3), R Foundation for Statistical Computing, Vienna, Austria).

### Ethical Considerations

The trial was approved by institutional review boards at the University of California, San Francisco and the Niger Ministry of Health.^25^ Written informed consent was obtained from parents or guardians of all participating children prior to enrollment and sample collection. The secondary analysis of stored samples for enteric fever serology was approved by the institutional review board at Massachusetts General Brigham.

## RESULTS

Samples from 1,417 children were analyzed: 424 from the baseline survey (2015), n=424 and 994 from the Year 5 survey (2020), following 10 rounds of biannual treatment (Figure 1). At baseline, the median age was 3.0 years (IQR 2.0–4.0) in both arms. Baseline characteristics were well balanced between arms, including sex distribution (57% male in placebo vs. 54% in azithromycin MDA), anemia status (moderate anemia in 45% vs. 43%), and malnutrition status (moderate/severe in 6.2% vs. 8.9%). Median HlyE IgA levels at baseline were similar between groups (3.4 K-ELISA units in both arms), as were HlyE IgG levels (15.5 vs. 14.8 K-ELISA units) (Table 1).

**Figure 1:**
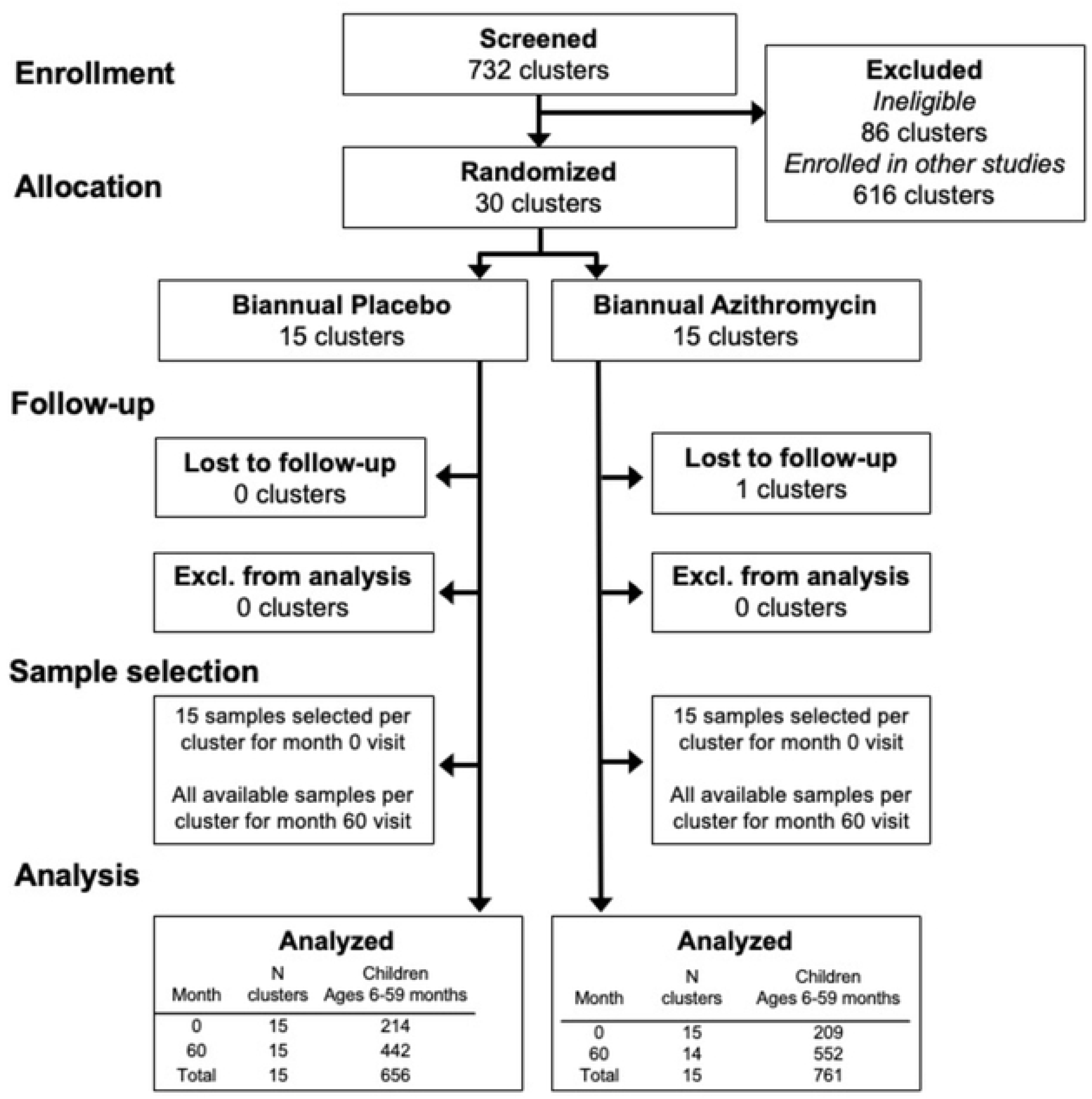
Participant flow diagram

**Table 1:** Baseline characteristics (pre-intervention) of participants included in the HlyE IgA/IgG analysis, by study arm.

| Characteristic | Study arm |  | N |
| --- | --- | --- | --- |
|  | Placebo<br>N = 209<sup>1</sup> | Azithromycin MDA<br>N = 214<sup>1</sup> |  |
| **Number of clusters** | 15 | 15 |  |
| **Children per cluster** | 14 (13, 14) | 15 (14, 15) |  |
| **Age, years** | 3.0 (2.0, 4.0) | 3.0 (2.0, 4.0) | 423 |
| **Sex** |  |  | 408 |
| Female | 89 (43%) | 91 (46%) |  |
| Male | 120 (57%) | 108 (54%) |  |
| Unknown | 0 | 15 |  |
| **Anemia status** |  |  | 408 |
| Normal | 43 (21%) | 51 (26%) |  |
| Mild | 61 (29%) | 54 (27%) |  |
| Moderate | 95 (45%) | 86 (43%) |  |
| Severe | 10 (4.8%) | 8 (4.0%) |  |
| Unknown | 0 | 15 |  |
| **Malnutrition status** |  |  | 423 |
| Moderate / Severe | 13 (6.2%) | 19 (8.9%) |  |
| Normal | 196 (94%) | 195 (91%) |  |
| **Mobile phone in household** |  |  | 408 |
| No | 205 (98%) | 194 (97%) |  |
| Yes | 4 (1.9%) | 5 (2.5%) |  |
| Unknown | 0 | 15 |  |
| **HlyE IgA** | 3.4 (2.4, 4.8) | 3.4 (2.2, 5.3) | 423 |
| **HlyE IgG** | 15.5 (10.2, 21.9) | 14.8 (9.6, 21.3) | 423 |

<sup>1</sup>Median (Q1, Q3); n (%)

Quantitative HlyE IgA and IgG antibody levels increased with age in both study arms at baseline and Year 5 (Figure 2). At baseline, HlyE IgA and IgG responses showed similar age-dependent increases in both the placebo and azithromycin arms, with overlapping smoothed trends. By Year 5, antibody levels were higher across all ages compared to baseline, but the age-specific trajectories remained similar between treatment groups for both isotypes. Population-level median antibody responses were comparable to confirmed enteric fever cases less than 300 days after infection (Supplemental Figure 1).

**Figure 2.**
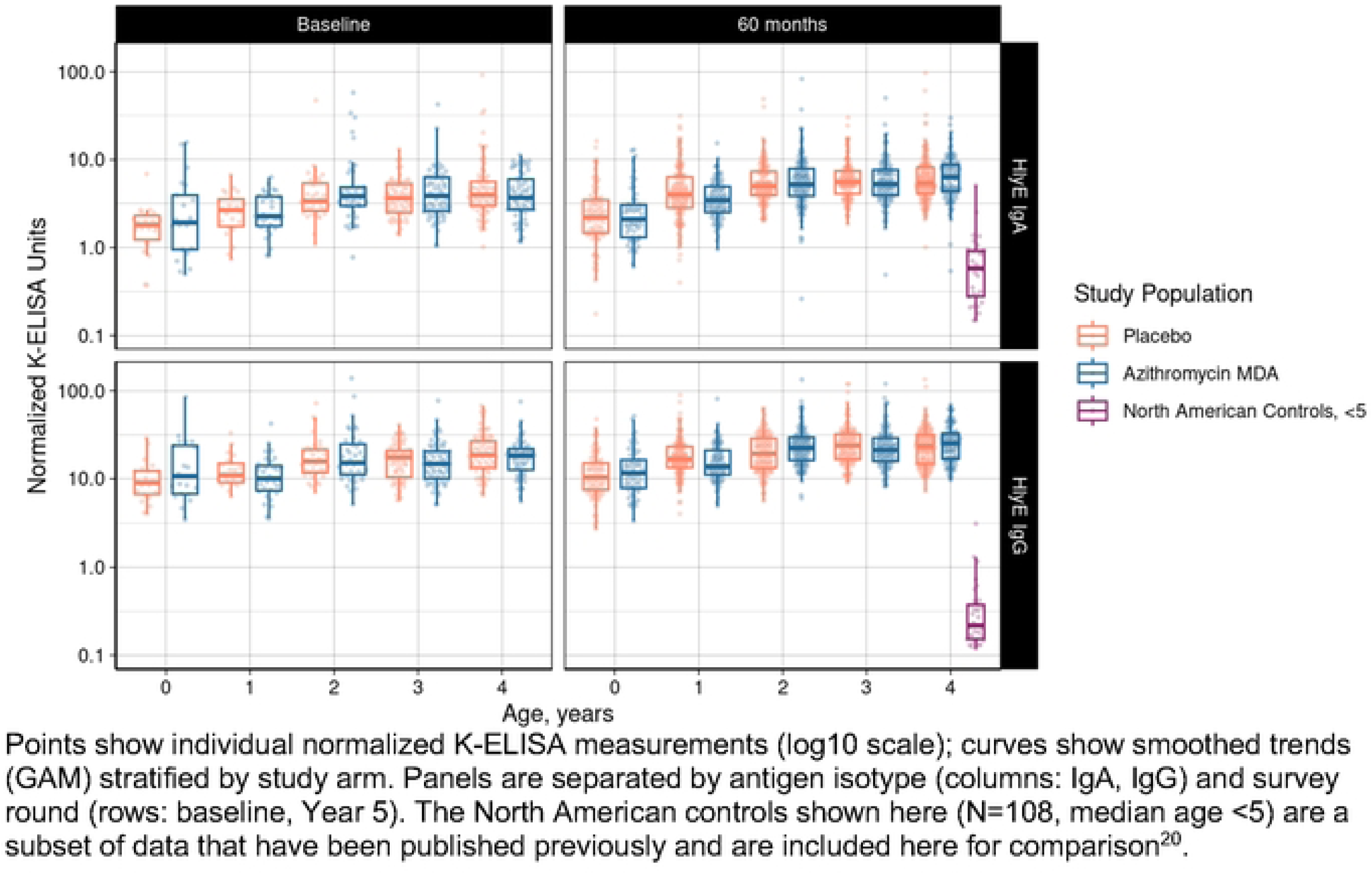
Age-specific HlyE IgA and IgG antibody levels by study arm at baseline and Year 5.

At baseline, HlyE IgA seroincidence varied significantly by age (Supplemental Table 1). Children under 1 year had the lowest seroincidence at 36.1 per 100 person-years (95% CI 14.3–91.5), which served as the reference group. Seroincidence increased with age, peaking at 99.9 per 100 person-years (95% CI 76.6–130.2) among 2-year-olds (p=0.003 vs. reference), then declining slightly to 73.8 per 100 person-years (95% CI 60.2–90.4) among 3-year-olds (p=0.045) and 69.8 per 100 person-years (95% CI 52.7–92.3) among 4-year-olds (p=0.089). Seroincidence did not differ significantly by sex: 63.1 per 100 person-years (95% CI 49.3–80.8) in females versus 78.2 per 100 person-years (95% CI 63.5–96.3) in males (p=0.190). Similarly, malnutrition status was not significantly associated with seroincidence. Children with normal nutritional status had a seroincidence of 70.7 per 100 person-years (95% CI 59.1–84.7), compared to 78.8 per 100 person-years (95% CI 46.7–133.0) in those with moderate malnutrition (p=0.713) and 142.1 per 100 person-years (95% CI 30.3–667.1) in those with severe malnutrition (p=0.525), though the latter estimate was imprecise due to small sample size (n=3).

Anemia status showed a trend toward higher seroincidence with increasing severity. Children with normal hemoglobin had a seroincidence of 57.0 per 100 person-years (95% CI 41.9–77.6), compared to 73.8 per 100 person-years (95% CI 59.0–92.4) with mild anemia (p=0.172), 73.0 per 100 person-years (95% CI 58.8–90.6) with moderate anemia (p=0.184), and 145.8 per 100 person-years (95% CI 76.6–277.4) with severe anemia (p=0.068).

Enteric fever seroincidence, estimated using HlyE IgA antibody kinetics, increased substantially from baseline to Year 5 in both study arms (Table 2). At baseline, overall seroincidence was 68.5 per 100 person-years (95% CI 58.0–80.7) in the placebo arm and 74.7 per 100 person-years (95% CI 56.5–98.7) in the azithromycin arm, with no significant difference between groups (difference 0.06; 95% CI −0.18 to 0.30). By Year 5, seroincidence had approximately doubled in both arms: 145.2 per 100 person-years (95% CI 132.3–159.3) in placebo and 135.2 per 100 person-years (95% CI 112.4–162.6) in azithromycin MDA arms. The difference between arms at Year 5 was not statistically significant (difference −0.10; 95% CI −0.38 to 0.18). The difference-in-differences estimate comparing the change from baseline to Year 5 between arms was -16.2 (95% CI: -53.1, 20.8).

**Table 2:** HlyE IgA seroincidence by study arm at baseline and Year 5.

| Survey round | Placebo<br>(N) | Placebo<br>seroincidence<br>(95% CI) | Azithromycin<br>MDA (N) | Azithromycin MDA<br>seroincidence (95%<br>CI) | Difference<br>(Azithro MDA –<br>Placebo, 95% CI) |
| --- | --- | --- | --- | --- | --- |
| **Age: Overall** |  |  |  |  |  |
| Baseline | 209 | 68.5 (58.0, 80.7) | 214 | 74.7 (56.5, 98.7) | 6.2 (-17.5, 29.9) |
| Year 5 | 552 | 145.2 (132.3,<br>159.3) | 442 | 135.2 (112.4, 162.6) | -10.0 (-38.3, 18.4) |
| Change<br>(Year 5 –<br>Baseline) |  | 76.7 (59.2, 94.3) |  | 60.6 (28.0, 93.1) | -16.2 (-53.1, 20.8) |
| **Age: <2 years** |  |  |  |  |  |
| Baseline | 44 | 33.7 (17.9, 63.3) | 52 | 64.0 (42.1, 97.4) | 30.3 (-4.0, 64.6) |
| Year 5 | 198 | 114.8 (99.2,<br>132.9) | 141 | 101.5 (80.0, 128.8) | -13.3 (-42.8, 16.1) |
| Change<br>(Year 5 –<br>Baseline) |  | 81.1 (54.0, 108.2) |  | 37.5 (1.3, 73.6) | -43.7 (-88.8, 1.5) |
| **Age: 2–4 years** |  |  |  |  |  |
| Baseline | 165 | 78.9 (64.0, 97.1) | 162 | 78.6 (58.3, 105.9) | -0.3 (-28.9, 28.3) |
| Year 5 | 354 | 191.4 (167.8,<br>218.4) | 301 | 172.7 (133.4, 223.5) | -18.8 (-70.0, 32.4) |
| Change<br>(Year 5 –<br>Baseline) |  | 112.6 (82.4,<br>142.7) |  | 94.1 (43.8, 144.4) | -18.5 (-77.1, 40.2) |

Seroincidence per 100 person-years (95% CI). 'Difference' is difference in seroincidence rates between Azithro MDA – Placebo at each timepoint; in the Change row, 'Difference' is the difference-in-differences (DiD). CIs use SE propagation assuming independence between survey rounds.

Age-stratified analyses revealed higher seroincidence among children aged 2–4 years compared to those under 2 years at both timepoints (Table 2, Figure 3). At baseline, seroincidence in children <2 years was 33.7 per 100 person-years (95% CI 17.9–63.3) in the placebo arm and 64.0 per 100 person-years (95% CI 42.1–97.4) in the azithromycin arm.Among children 2–4 years, baseline seroincidence was 78.9 per 100 person-years (95% CI 64.0–97.1) in the placebo arm and 78.6 per 100 person-years (95% CI 58.3–105.9) in the azithromycin arm.

**Figure 3:**
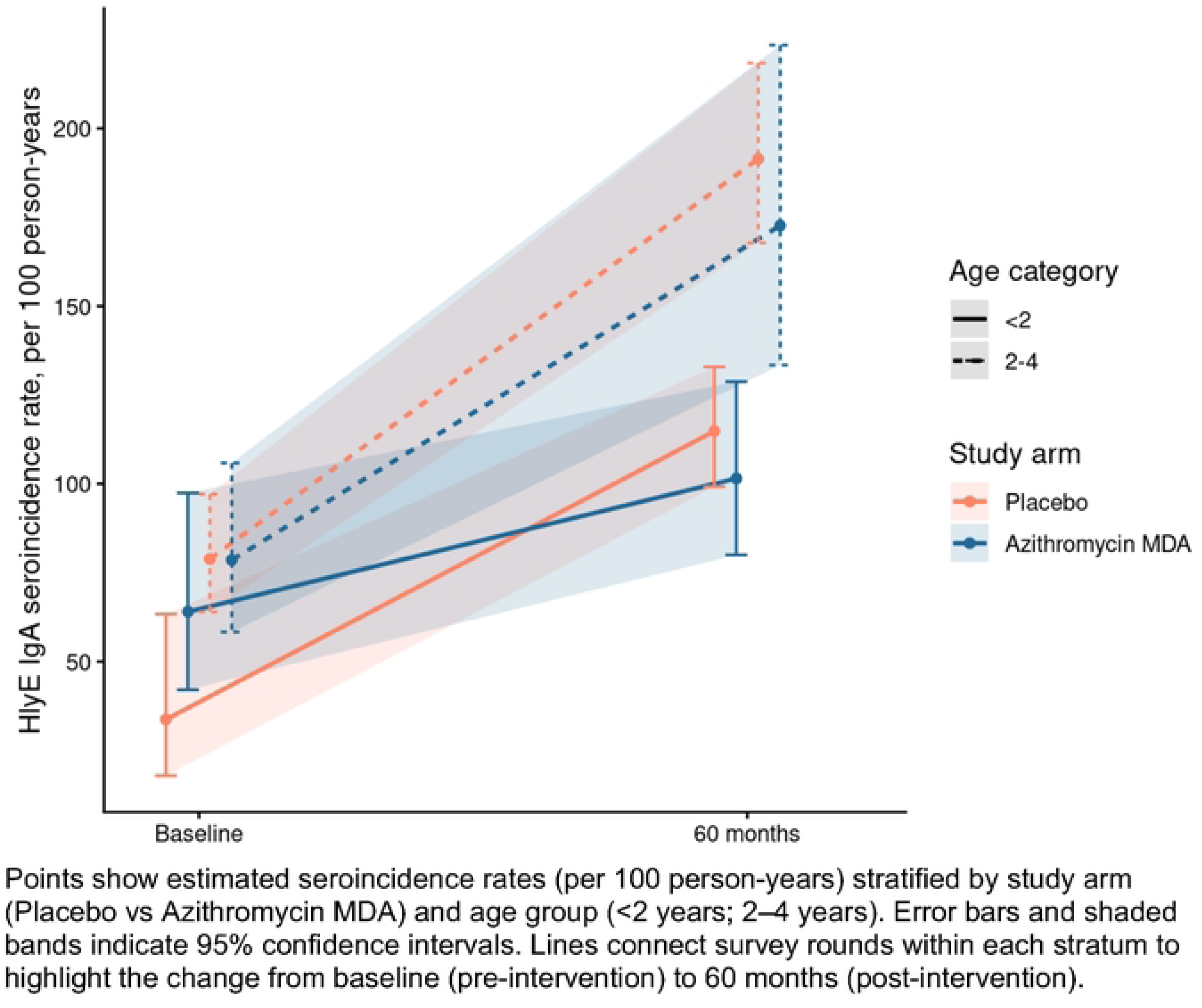
HlyE IgA seroincidence estimates at baseline and 60 months by study arm and age category.

By Year 5, seroincidence in children <2 years increased to 114.8 per 100 person-years (95% CI 99.2–132.9) in placebo and 101.5 per 100 person-years (95% CI 80.0–128.8) in azithromycin. The change from baseline to Year 5 was 81.1 per 100 person-years (95% CI 80.9–81.4) in placebo compared to 37.5 per 100 person-years (95% CI 37.1–37.9) in azithromycin, yielding a difference-in-differences of −43.7 per 100 person-years (95% CI −44.1 to −43.2). Among children aged 2–4 years, seroincidence at Year 5 was 191.4 per 100 person-years (95% CI 167.8–218.4) in placebo and 172.7 per 100 person-years (95% CI 133.4–223.5) in azithromycin. The change from baseline to Year 5 was 112.6 per 100 person-years (95% CI 112.3–112.9) in placebo versus 94.1 per 100 person-years (95% CI 93.6–94.6) in azithromycin, with a difference-in-differences of −18.5 per 100 person-years (95% CI −19.1 to −17.9).

Substantial heterogeneity in seroincidence was observed across clusters (Figure 4). At baseline, cluster-level HlyE IgA seroincidence ranged widely within both arms. By Year 5, most clusters showed increased seroincidence compared to baseline, with similar patterns of increase in placebo and azithromycin communities. Cluster-level seroincidence at baseline and Year 5 was modestly correlated over time (r=0.40).

**Figure 4.**
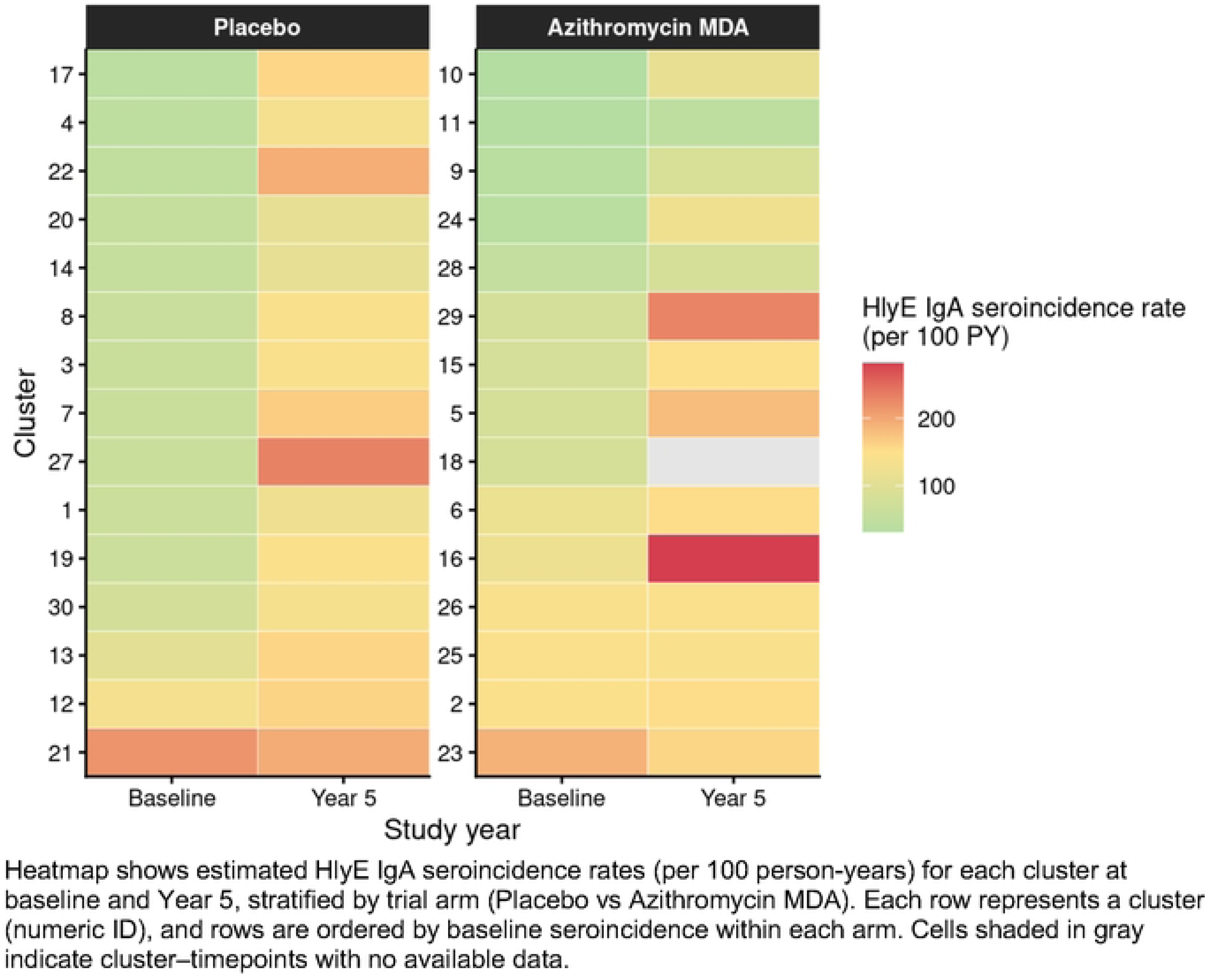
Cluster-level HlyE IgA seroincidence at baseline and Year 5 by study arm. Heatmap shows estimated HlyE IgA seroincidence rates (per 100 person-years) for each cluster at baseline and Year 5, stratified by trial arm (Placebo vs Azithromycin MDA). Each row represents a cluster (numeric ID), and rows are ordered by baseline seroincidence within each arm. Cells shaded in gray indicate cluster-timepoints with no available data.

## DISCUSSION

This study demonstrates an exceptionally high burden of enteric fever among young children in rural Niger, a setting without blood culture surveillance. Using validated serological methods, we estimated HlyE IgA seroincidence rates of 68.5 per 100 person-years at baseline (2015), increasing to 145.2 and 135.2 per 100 person-years in placebo and azithromycin communities, respectively, by Year 5 (2020). Biannual azithromycin MDA did not demonstrate a significant overall effect on enteric fever seroincidence. However, children under 2 years in the intervention group had the smallest rise in enteric fever seroincidence, suggesting a potential protective effect.

The seroincidence rates observed in rural Niger are among the highest reported globally using these serological methods. In comparison, HlyE seroincidence estimates have been reported in South Asia ranging from 6.6 per 100 person-years in Kavrepalanchok, Nepal to 58.5 per 100 person-years in Dhaka, Bangladesh among children under 5 years^20^. Seroincidence in Juba, South Sudan—another setting without blood culture surveillance—was estimated at 42.5 per 100 person-years among children 1 to 3 years old^23^. The Niger estimates substantially exceed these values, consistent with indirect evidence from surgical case series documenting exceptionally high rates of typhoid intestinal perforation in the country^7,8^.

The reasons for the increase in enteric fever seroincidence between 2015 and 2020 are not clear. Several factors may have contributed to the rise observed in both study arms. One possible explanation is environmental and infrastructural changes occurring in the Dosso region during this period. A recent analysis of the Dosso region documented rapid settlement expansion into flood-prone areas between 2004 and 2019, with approximately 80% of settlements affected by pluvial flooding^30^. Previous studies have linked flooding to increased typhoid risk through contamination of water sources and disruption of sanitation systems^31–33^. However, because we did not measure flooding, water quality, sanitation, infrastructure, or other potential drivers of infection in the study communities, these explanations remain speculative. Additional studies incorporating environmental and WASH indicators would be needed to better understand the factors underlying the increase in seroincidence.

The lack of a significant overall reduction in enteric fever seroincidence in the azithromycin MDA may reflect the very high force of infection in this setting. Azithromycin has a half-life of approximately 68 hours, and while it is an effective treatment for uncomplicated enteric fever, the protective window may be insufficient to substantially reduce cumulative exposure over a 6-month dosing interval when infection intensity is near-continuous. A serological substudy in the same MORDOR Niger communities found that biannual azithromycin MDA reduced *Campylobacter* spp. force of infection but showed no significant differences between treatment groups for other bacterial pathogens, including *Salmonella*, against a backdrop of high transmission^26^. Among children under 2 years, the smaller increase in seroincidence in azithromycin communities compared to placebo suggests a potential age-specific effect that merits further investigation.

In contrast to South Asian cohorts where HlyE IgG and IgA seroincidence estimates have been comparable^20^, HlyE IgG seroincidence estimates in Niger were substantially higher than those derived from IgA. This pattern may reflect the cumulative effect of repeated subclinical exposures in a very high burden setting, where IgG responses are boosted and maintained, while IgA responses are more transient and appear less influenced by repeat infections. Similar patterns have been observed in other African settings, where anti-HlyE IgG responses appear elevated relative to anti-HlyE IgA compared with populations in Asia^29^. A recent expert consultation convened by the Typhoid Vaccine Acceleration Consortium (TyVAC) concluded that anti-HlyE IgA should be prioritized for seroincidence estimation because its decay kinetics appear more consistent across settings and less sensitive to reinfection-driven boosting^29^.

These findings support the use of IgA-based seroincidence estimates for comparing infection intensity across populations and suggest that IgA may provide a more stable and generalizable measure of enteric fever burden in high-transmission settings where IgG kinetics may differ from those observed in lower-burden populations.

These findings have direct implications for typhoid prevention policy. The WHO recommends TCV introduction in countries with high typhoid fever incidence (≥100 per 100,000 person-years) or high prevalence of antimicrobial-resistant *S.* Typhi^34^. The seroincidence rates observed in rural Niger far exceed this threshold. Niger introduced TCV in late 2025, reaching children aged 1-19 years through a national campaign. The present findings provide additional evidence supporting the recent introduction of TCV in Niger and underscore the importance of monitoring vaccine impact in this high-burden setting.

This study has several limitations. The seroincidence estimation method relies on antibody decay parameters derived from blood culture-confirmed enteric fever patients in South Asia, which may not fully capture antibody dynamics in African populations with different exposure histories. Because the baseline and Year 5 surveys sampled different children, observed differences may reflect temporal changes in transmission as well as changes in the underlying population sampled. The study population was limited to children aged 1–59 months, and infection patterns in older age groups remain unknown. Confidence intervals for difference-in-differences estimates assumed independence between survey rounds and may therefore be conservative given the modest within-cluster correlation in seroincidence over time. The samples were tested between 4 and 9 years after sample collection, depending on the study visit. It’s possible that antibodies degraded overtime and during shipment despite temperature-controlled storage conditions; this possible degradation would result in underestimated seroincidence. Finally, HlyE is expressed by both *S.* Typhi and *S.* Paratyphi A, precluding distinction between these serovars, although paratyphoid fever is rare in African settings^35^.

In conclusion, this study documents a high burden of enteric fever among young children in rural Niger and demonstrates the utility of serological surveillance methods for estimating infection intensity in settings without blood culture infrastructure. Biannual azithromycin MDA did not significantly reduce overall seroincidence, although a potential protective effect among the youngest children warrants further investigation. These data support the recent introduction of typhoid conjugate vaccine in Niger and highlight the need for continued surveillance to assess vaccine impact.

## Data Availability

De-identified data will be available upon publication in OSF

https://osf.io/ne8pc/overview

## REFERENCES

1 Stanaway JD, Reiner RC, Blacker BF, et al. The global burden of typhoid and paratyphoid fevers: a systematic analysis for the Global Burden of Disease Study 2017. Lancet Infect Dis 2019; 19: 369–81.

2 Andrews JR, Barkume C, Yu AT, et al. Integrating Facility-Based Surveillance With Healthcare Utilization Surveys to Estimate Enteric Fever Incidence: Methods and Challenges. J Infect Dis 2018; 218: S268–76.

3 Mogasale V, Ramani E, Mogasale VV, Park J. What proportion of Salmonella Typhi cases are detected by blood culture? A systematic literature review. Ann Clin Microbiol Antimicrob 2016; 15: 32.

4 Carey ME, MacWright WR, Im J, et al. The Surveillance for Enteric Fever in Asia Project (SEAP), Severe Typhoid Fever Surveillance in Africa (SETA), Surveillance of Enteric Fever in India (SEFI), and Strategic Typhoid Alliance Across Africa and Asia (STRATAA) Population-based Enteric Fever Studies: A Review of Methodological Similarities and Differences. Clin Infect Dis 2020; 71: S102–10.

5 von Kalckreuth V, Konings F, Aaby P, et al. The Typhoid Fever Surveillance in Africa Program (TSAP): Clinical, Diagnostic, and Epidemiological Methodologies. Clin Infect Dis Off Publ Infect Dis Soc Am 2016; 62 **Suppl 1**: S9–16.

6 Troeger C, Forouzanfar M, Rao PC, et al. Estimates of global, regional, and national morbidity, mortality, and aetiologies of diarrhoeal diseases: a systematic analysis for the Global Burden of Disease Study 2015. Lancet Infect Dis 2017; 17: 909–48.

7 Sukri L, Banza A, Shafer K, Sanoussi Y, Neuzil KM, Sani R. Typhoid intestinal perforation in Francophone Africa, a scoping review. PLOS Glob Public Health 2024; 4: e0003056.

8 Adamou H, Amadou Magagi I, Adakal O, et al. Le fardeau de la perforation typhique de l’intestin grêle au Niger. The burden of typhoid perforation of the small intestine in Niger. 2021; 1: 131–9.

9 Qazi SH, Yousafzai MT, Saddal NS, et al. Burden of Ileal Perforations Among Surgical Patients Admitted in Tertiary Care Hospitals of Three Asian countries: Surveillance of Enteric Fever in Asia Project (SEAP), September 2016-September 2019. Clin Infect Dis Off Publ Infect Dis Soc Am 2020; 71: S232–8.

10 Murthy S, Hagedoorn NN, Faigan S, Rathan MD, Marchello CS, Crump JA. Complications and mortality of typhoid fever: an updated global systematic review and meta-analysis. Lancet Infect Dis 2025; : S1473-3099(25)00551-1.

11 Kuehn R, Rahden P, Hussain HS, et al. Enteric (typhoid and paratyphoid) fever. Lancet Lond Engl 2025; 406: 1283–94.

12 Keenan JD, Arzika AM, Maliki R, et al. Longer-Term Assessment of Azithromycin for Reducing Childhood Mortality in Africa. N Engl J Med 2019; 380: 2207–14.

13 Keenan JD, Bailey RL, West SK, et al. Azithromycin to reduce childhood mortality in sub-Saharan Africa. N Engl J Med 2018; 378: 1583–92.

14 Sié A, Ouattara M, Bountogo M, et al. Mass Azithromycin Distribution and Cause-Specific Mortality among Children Ages 1-59 Months Old: A Secondary Analysis of a Cluster-Randomized Controlled Trial. Am J Trop Med Hyg 2025; 114: 85–91.

15 Oldenburg CE, Ouattara M, Bountogo M, et al. Mass Azithromycin Distribution to Prevent Child Mortality in Burkina Faso: The CHAT Randomized Clinical Trial. JAMA 2024; 331: 482– 90.

16 Hooda Y, Sajib MSI, Rahman H, et al. Molecular mechanism of azithromycin resistance among typhoidal Salmonella strains in Bangladesh identified through passive pediatric surveillance. PLoS Negl Trop Dis 2019; 13: e0007868.

17 Sajib MSI, Tanmoy AM, Hooda Y, et al. Tracking the Emergence of Azithromycin Resistance in Multiple Genotypes of Typhoidal Salmonella. mBio 2021; 12: e03481–20.

18 Carey ME, Jain R, Yousuf M, et al. Spontaneous Emergence of Azithromycin Resistance in Independent Lineages of Salmonella Typhi in Northern India. Clin Infect Dis Off Publ Infect Dis Soc Am 2021; 72: e120–7.

19 Qamar FN, Yousafzai MT, Dehraj IF, et al. Antimicrobial Resistance in Typhoidal Salmonella: Surveillance for Enteric Fever in Asia Project, 2016-2019. Clin Infect Dis Off Publ Infect Dis Soc Am 2020; 71: S276–84.

20 Aiemjoy K, Seidman JC, Saha S, et al. Estimating typhoid incidence from community-based serosurveys: a multicohort study. Lancet Microbe 2022; 3: e578–87.

21 Aiemjoy K, Seidman JC, Charles RC, Andrews JR. Seroepidemiology for Enteric Fever: Emerging Approaches and Opportunities. Open Forum Infect Dis 2023; 10: S21–5.

22 Charles RC, Sheikh A, Krastins B, et al. Characterization of Anti-Salmonella enterica Serotype Typhi Antibody Responses in Bacteremic Bangladeshi Patients by an Immunoaffinity Proteomics-Based Technology. Clin Vaccine Immunol CVI 2010; 17: 1188– 95.

23 Aiemjoy K, Rumunu J, Hassen JJ, et al. Seroincidence of Enteric Fever, Juba, South Sudan. Emerg Infect Dis 2022; 28: 2316–20.

24 Khan A, Kamenskaya P, Rezende I, et al. Seroincidence Rate of Typhoidal Salmonella in Children, Kenya, 2017–2018 - Volume 32, Number 3—March 2026 - Emerging Infectious Diseases journal - CDC. DOI:10.3201/eid3203.250469.

25 Keenan JD, Bailey RL, West SK, et al. Azithromycin to Reduce Childhood Mortality in Sub-Saharan Africa. N Engl J Med 2018; 378: 1583–92.

26 Arzika AM, Maliki R, Goodhew EB, et al. Effect of biannual azithromycin distribution on antibody responses to malaria, bacterial, and protozoan pathogens in Niger. Nat Commun 2022; 13: 976.

27 Teunis PFM, van Eijkeren JCH, de Graaf WF, Marinović AB, Kretzschmar MEE. Linking the seroresponse to infection to within-host heterogeneity in antibody production. Epidemics 2016; 16: 33–9.

28 Teunis PFM, van Eijkeren JCH, Ang CW, et al. Biomarker dynamics: estimating infection rates from serological data. Stat Med 2012; 31: 2240–8.

29 Laurens M, Ackah E, Agyapong F, et al. Typhoid Seroepidemiology for TCV Decision Making Meeting Report of the 18 July 2025 Expert Consultation [version 1]. VeriXiv 2026; 3. DOI:10.12688/verixiv.3487.1.

30 Tiepolo M, Galligari A. Urban expansion-flood damage nexus: Evidence from the Dosso Region, Niger. Land Use Policy 2021; 108: 105547.

31 Balikuddembe JK, Luo X, Di B, et al. Association of typhoid fever with floods under climate variability in 82 Belt and Road Initiative countries (2000-2021): A mixed-effects model and implications for water and sanitation infrastructure. Sci Total Environ 2025; 1002: 180629.

32 Okyere PB, Twumasi-Ankrah S, Newton S, et al. Risk Factors for Typhoid Fever: Systematic Review. JMIR Public Health Surveill 2025; 11: e67544.

33 Liu Z, Lao J, Zhang Y, et al. Association between floods and typhoid fever in Yongzhou, China: Effects and vulnerable groups. Environ Res 2018; 167: 718–24.

34 World Health Organization. Typhoid vaccines: WHO position paper, March 2018 – Recommendations. Vaccine 2019; 37: 214–6.

35 Marks F, von Kalckreuth V, Aaby P, et al. Incidence of invasive salmonella disease in sub-Saharan Africa: a multicentre population-based surveillance study. Lancet Glob Health 2017; 5: e310–23.

